# PADI4 minor haplotype as risk factor for excessive NET formation and associated wound healing disorders in diabetes mellitus

**DOI:** 10.1101/2024.11.28.24318124

**Authors:** Sabrina Ehnert, Philipp Hemmann, Christoph Ihle, Caren Linnemann, Jonas Mück, Panagiota-Georgia Anastasiou, Ralf Lobmann, Gunnar Blumenstock, Stefan Pscherer, Andreas Fritsche, Heiko Baumgartner, Tina Histing, Mika F. Rollmann, Andreas K. Nussler

**Affiliations:** Siegfried Weller Research Institute; BG Unfallklinik Tübingen; Department of Trauma and Reconstructive Surgery; University of Tübingen; Schnarrenbergstr. 95; D-72076 Tübingen; Germany; Clinic for Endocrinology; Diabetology and Geriatric Medicine; Center for Internal Medicine; Klinikum der Landeshauptstadt Stuttgart gKAöR, Katharinenhospital; Kriegsbergstrasse 60, D-70174 Stuttgart; Germany; Institute for Clinical Epidemiology and Applied Biometry; University of Tübingen; Silcherstr. 5; D-72076 Tübingen; Germany; Department of Internal Medicine III, Sophien- and Hufeland-Hospital, Henry-van-de-Velde-Straße 2, D-99425 Weimar, Germany; Department of Internal Medicine, Division of Diabetology, Endocrinology, and Nephrology, Eberhard Karls University Tübingen, Otfried-Müller-Straße 10, D-72076 Tübingen, Germany

**Keywords:** Diabetes, Wound Healing, Tissue Infection, Neutrophil Extracellular Traps, PADI4 haplotype

## Abstract

Diabetes mellitus (DM) is associated with impaired wound healing, partly driven by excessive neutrophil extracellular trap (NET) formation mediated by peptidyl-arginine deiminase 4 (PADI4). While circulating NET markers predict poor healing outcomes, they likely reflect established tissue damage and offer limited opportunity for early intervention. We therefore investigated the association between PADI4 haplotypes, PADI4 expression, NET formation, and clinical outcomes, namely infections and delayed wound and bone healing, in 687 surgical patients (44.7% with DM). Pre-surgical PADI4 expression was 9.4-fold higher in patients with DM, particularly in those who developed wound healing complications. Neutrophils carrying the PADI4 minor haplotype showed increased PADI4 mRNA and protein expression and produced larger quantities of NETs more rapidly than those with the major haplotype. Clinically, patients with DM carrying the minor haplotype had the highest rates of delayed wound healing and infections. Together, these results demonstrate that PADI4 genetic variation influences neutrophil behavior and clinical outcomes. PADI4 haplotyping may provide a clinically actionable biomarker to identify patients with DM at high risk for wound healing complications and guide early preventive strategies.

**Graphical Abstract:** 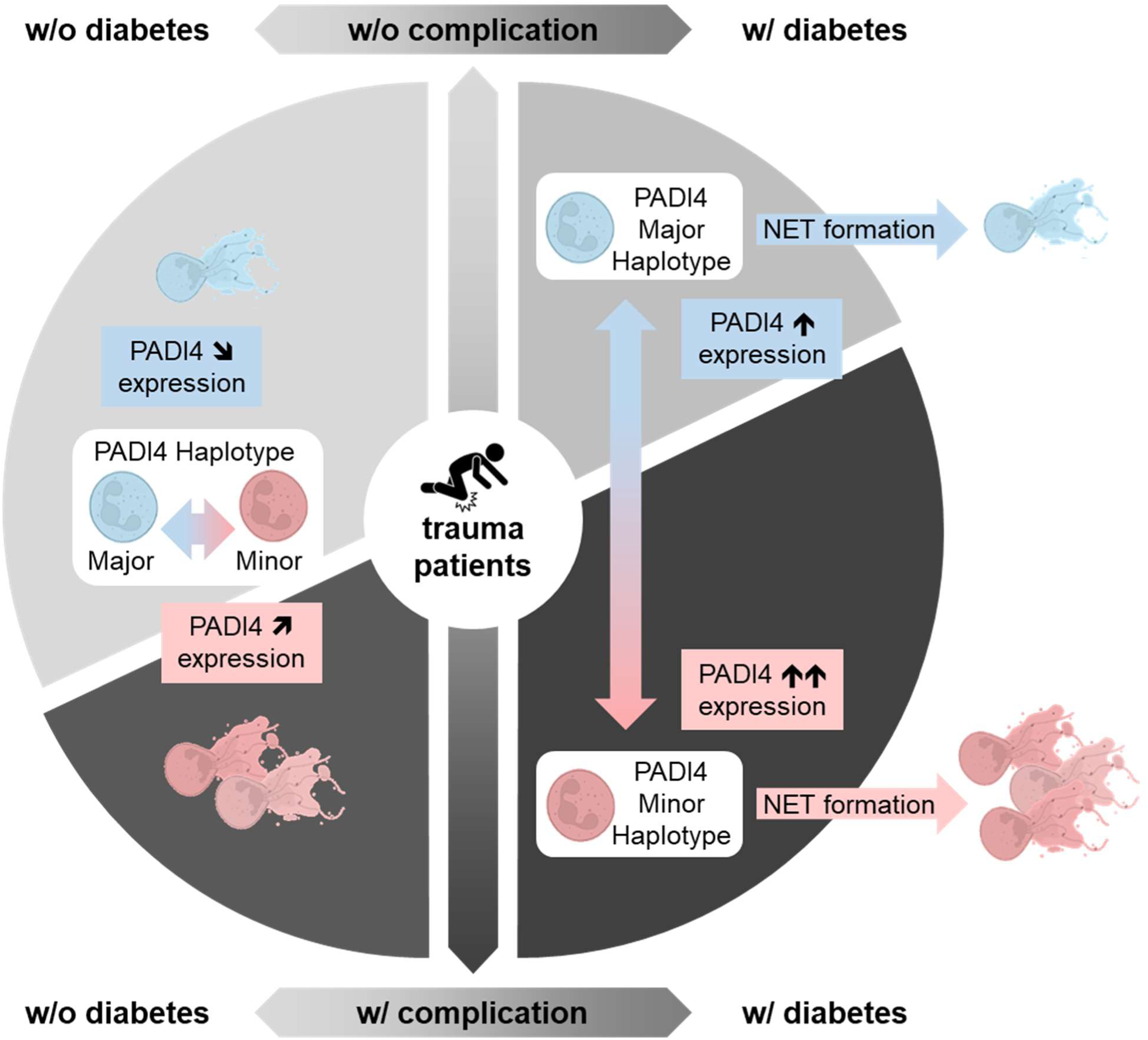

## Introduction

A constantly aging society and sedentary lifestyle made Diabetes mellitus (DM), especially DM type 2, the disease with the fastest growing incidence and prevalence worldwide. Based on reports from the world health organization, DM is a major cause of blindness, kidney failure, heart attacks, or stroke. Likewise, it is indisputable that DM affects the quality of bones and surrounding soft tissues, increasing the risk of fractures and associated healing disorders, *e.g.*, infections and/or delayed bone and wound healing. Approximately every tenth develops a foot ulcer, diabetic foot syndrome, or charcot-osteoarthropathy within the first 5 years of the disease. Of these roughly 20% will have amputations of the lower limbs in the following 5 years, making DM the leading cause of non-traumatic amputations worldwide (*1*). The resulting treatment costs exceed by far the costs for treating the underlying disease (*2*), underlining the need to develop strategies to prevent these diabetes-related complications.

For most complications, it is generally believed that patients with poor glycemic control and longer disease duration are at highest risk. Considering diabetes-related complications of the kidneys, liver, nerves and cardiovascular system, additional risk factors have been identified (*3, 4*). It is assumed that certain anti-diabetic drugs affect bone metabolism and thus influence the fracture risk (*6*). For diabetes-associated complications following orthopedic or trauma surgery, information on risk factors, and consequently treatment recommendations, are yet limited (*5*). In diabetic mice, a massive invasion of neutrophils into wounds was described. Within the wounds, neutrophils were activated to form so-called neutrophil extracellular traps (NETs) (*7*), an unspecific defense mechanism where neutrophils release their DNA together with citrullinated histones and anti-microbial peptides in order to bind and neutralize pathogens (*8*). In wounds of diabetic mice, however, an accumulation of NETs was observed, which impaired the wound healing (*7*), either indirectly by stimulating local inflammation (*9*) or directly by acting as damage-associated molecular patterns harming the surrounding tissue (*10*). These findings in mice have been reproduced in different laboratories ever since (*11–13*). In humans, circulating NETs markers, *e.g.*, neutrophil elastase (ELANE), myeloperoxidase (MPO), citrullinated histones H3 (citH3), cell-free (cf) DNA, and nucleosomes, were found to be increased in patients with DM, especially when having foot ulcers (DFUs) - for review see (*14*). The levels of ELANE and citH3 were negatively associated with the DFUs’ spontaneous healing potential while relating to the amputation rate (*12, 15*). These findings emphasize the important role of NET accumulation for impaired wound healing in patients with DM, and thus provide opportunities for the development of novel treatment strategies. However, when considering that the accumulated NETs may severely damage the surrounding tissue, measuring circulating NET markers may not allow enough time to adjust the treatment in the clinical setting. In order to gain time for treatment a biomarker that indicates the potential of neutrophils to form NETs would be required.

Overexpression of peptidyl arginine deiminase 4 (PADI4) in neutrophils was identified to be responsible for the overshooting NET formation in wounds of diabetic mice (*7*). PADI4 is known to convert arginine residues to citrulline in proteins – in case of NETosis mainly in histones, inducing nuclear decondensation (*16*). Even before NETs were first described, a dysregulated PADI4 activity, was related to abnormal citrullination of auto-antibodies in patients with rheumatoid arthritis (RA) - for review see (*17*). The link between PADI4 and onset and progression of RA was strengthened when a genome wide association study identified a RA-associated PADI4 haplotype. This PADI4 functional haplotype consists of 3 single nucleotide polymorphisms (SNPs: rs11203366, rs11203367, rs874881), all located on the first 2 exons of the PADI4 gene, resulting in 3 amino acid substitutions in the N-terminal region of the PADI4 enzyme, affecting its structure (*18*). Special about this PADI4 haplotype is its high minor allele frequency, due to which approx. 22% of the worldwide population are homozygous for the minor variant (mm; Figure 2A). Recently, other SNPs in the PADI4 gene were proposed as potential risk loci in patients with rheumatoid arthritis, *e.g.*, SNP rs2301888 (*19*), rs2240336, and rs766449 (*20*), however, these SNPs do not belong to the described haplotype (R^2^ < 0.37, < 0.22, < 0.59, respectively) and failed statistical significance for an European population (*19, 20*), which is the expected majority in this study cohort.

Although increased PADI4 activity was associated with excessive NET formation in non-healing diabetics wounds (*14*), no such relationships between the PADI4 haplotype and the healing potential of diabetic wounds or DFUs was shown so far. Based on the mentioned literature it was hypothesized that the PADI4 haplotype influences the neutrophils response to NET-stimuli and that this relates to the clinical outcome of patients with DM undergoing trauma and/or orthopedic surgery. Therefore, this study aimed to investigate the relationship between PADI4 haplotypes, pre-surgical neutrophilic PADI4 expression, and clinical outcomes – specifically delayed wound healing, tissue infections, and delayed bone healing – in patients (with and without diabetes) who underwent surgery at a level 1 trauma center. Additionally, the study sought to determine whether PADI4 haplotypes influence the potential of neutrophils to form NETs.

## Results

### Patients with DM have higher complication rates than patients without DM

With the overall aim that the patients without DM fit those with DM regarding age, gender, and BMI, 725 patients qualified for our study. 38 patients were excluded from the study because they refused to participate or blood sampling was not possible. Of the remaining 687 patients, 380 were assigned for the control group (without DM) and 307 were assigned for the DM group, based on their medical history, random blood glucose levels, and/or HbA1C values. The clinical data were independently documented by two clinicians without knowing the laboratory data. The development of infections, delayed wound healing, or delayed bone healing following primary interventions was counted as complications. Revision surgeries were counted as complications when fitting into the above defined categories – from these patients PADI4 haplotype was determined but not neutrophilic PADI4 gene expression. Following this classification, the control group contained 245 patients (112 males and 133 females) without complications and 135 patients (90 males and 45 females) with complications. Similarly, the DM group contained 112 patients (61 males and 51 females) without complications and 195 patients (129 males and 66 females) with complications. Interestingly, in the latter there was no significant difference in HbA1C values of patients with (7.8 ± 1.8% / 4.7 – 14.0) and without complications (9.0 ± 3.2% / 5.3 – 17.6; Figure 1A). The VENN diagrams show frequent co-occurrence of 2 or 3 of the rated complications, especially in patients with DM (Figure 1B). The age distribution revealed that patients without DM (w/o: 59.4 ± 13.7 years / w/: 57.6 ± 14.9 years) were on average 6 years younger than patients with DM (w/o: 63.5 ± 12.9 years / w/: 64.7 ± 11.4 years), regardless of the occurrence of a complication (Figure 1C). Similarly, the body mass index (BMI) was increased in patients with DM and complications (30.7 ± 6.1 kg/m^2^) as compared to the respective controls (28.4 ± 5.8 kg/m^2^; Figure 1D). In diabetic mice increased neutrophilic PADI4 expression was associated with impaired wound healing (*7*). Therefore, PADI4 expression was determined in neutrophils, isolated prior to primary surgery. Basal PADI4 expression was significantly increased (1.9 ± 2.5 *vs.* 17.7 ± 45.6; *p* < 0.0001) in neutrophils from patients with DM when compared to matched patients without DM (Figure 1E).

**Figure 1:**
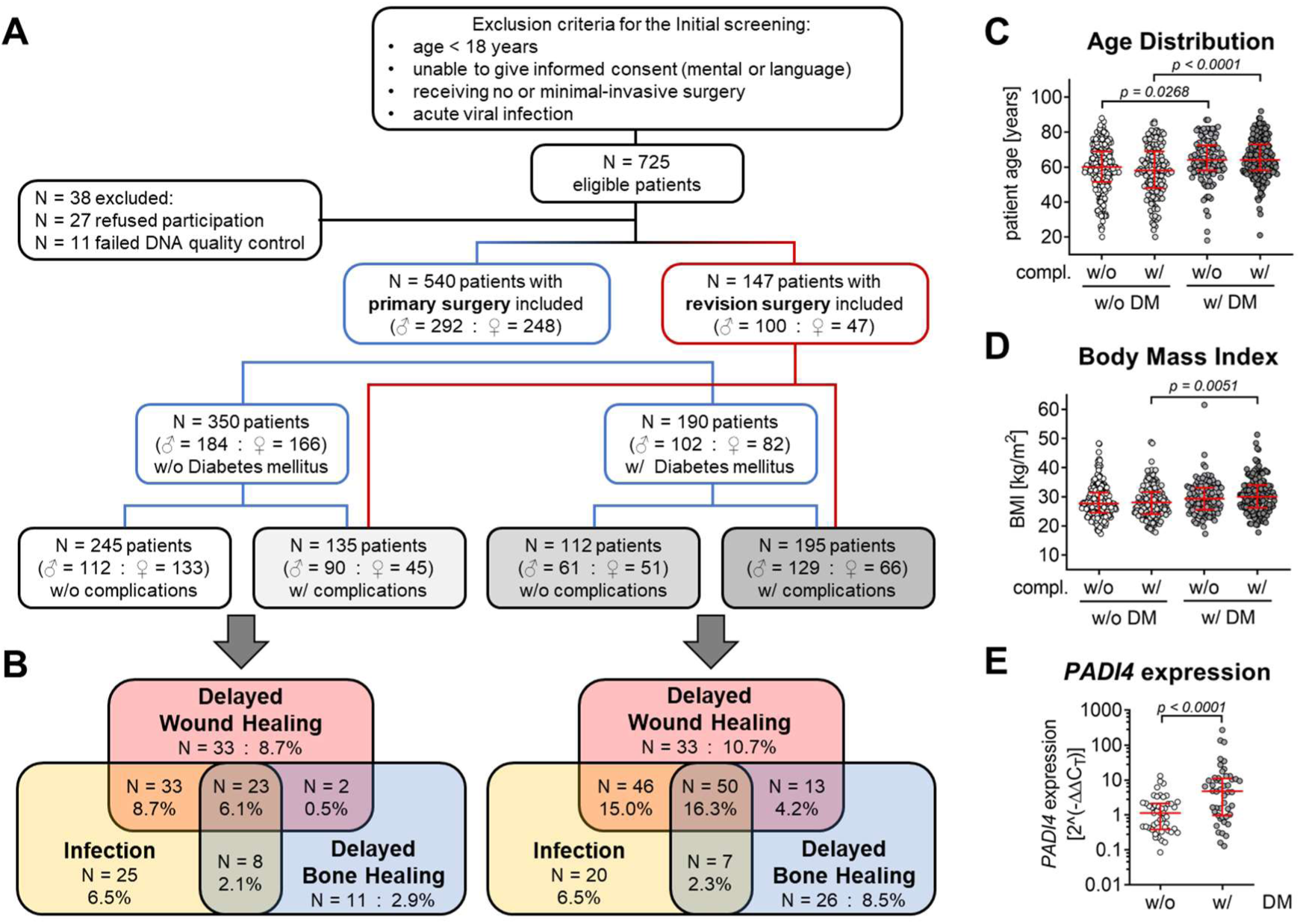
Overview on the patients’ enrolment in this study. (A) Patient selection process according to the STREGA guidelines, including patient assignment into the four study groups: patients without diabetes mellitus (Ctrl) and with diabetes mellitus (DM), each with (w/) and without (w/o) complications. Disturbed wound healing, delayed bone healing, and post-surgical infections were counted as complications – when developed within 6 months after a primary intervention. (B) The distribution of the complications is represented as VENN diagrams for patients with and without DM. For each group (C) the patients’ age and (D) the individual body mass index (BMI) were obtained. (E) in addition, pre-surgical (primary intervention) PADI4 expression in circulating neutrophils was determined by quantitative real-time PCR in age and gender matched patients (each N = 48). Data are displayed in scatter plots with median and inter-quartile range. Groups were compared by (C&D) ordinary one-way ANOVA followed by Tukey’s multiple comparison test or (E) Mann-Whitney test. A *p*<0.05 was considered statistically significant and is shown in the graphs.

### PADI4 genotype distribution is independent of the patients’ sex, age, BMI, or prevalence for diabetes mellitus

All donors were characterized for PADI4 SNPs rs11203366, rs11203367, rs874881 by amplification-refractory mutation system polymerase chain reaction (ARMS-PCR). The haplotype frequency within the study cohort (51.2% A·C·C / 43.1% G·T·G / 5.7% other combinations) was comparable to the reference data available from the 1000 genomes project. Based on these data, 194 patients (28.2%) were identified to be homozygous for the major variant (MM) of PADI4, 116 patients (16.9%) were identified to be homozygous for the minor variant (mm) of PADI4, and the remaining 377 patients (54.9%) were identified to be heterozygous (Mm) of PADI4 (Figure 2A-C). Subgroup analyses revealed, that the PADI4 genotype distribution was independent of the patients’ age (Figure 2D), BMI (Figure 2E), sex (Figure 2F), or prevalence for diabetes mellitus (Figure 2G).

**Figure 2:**
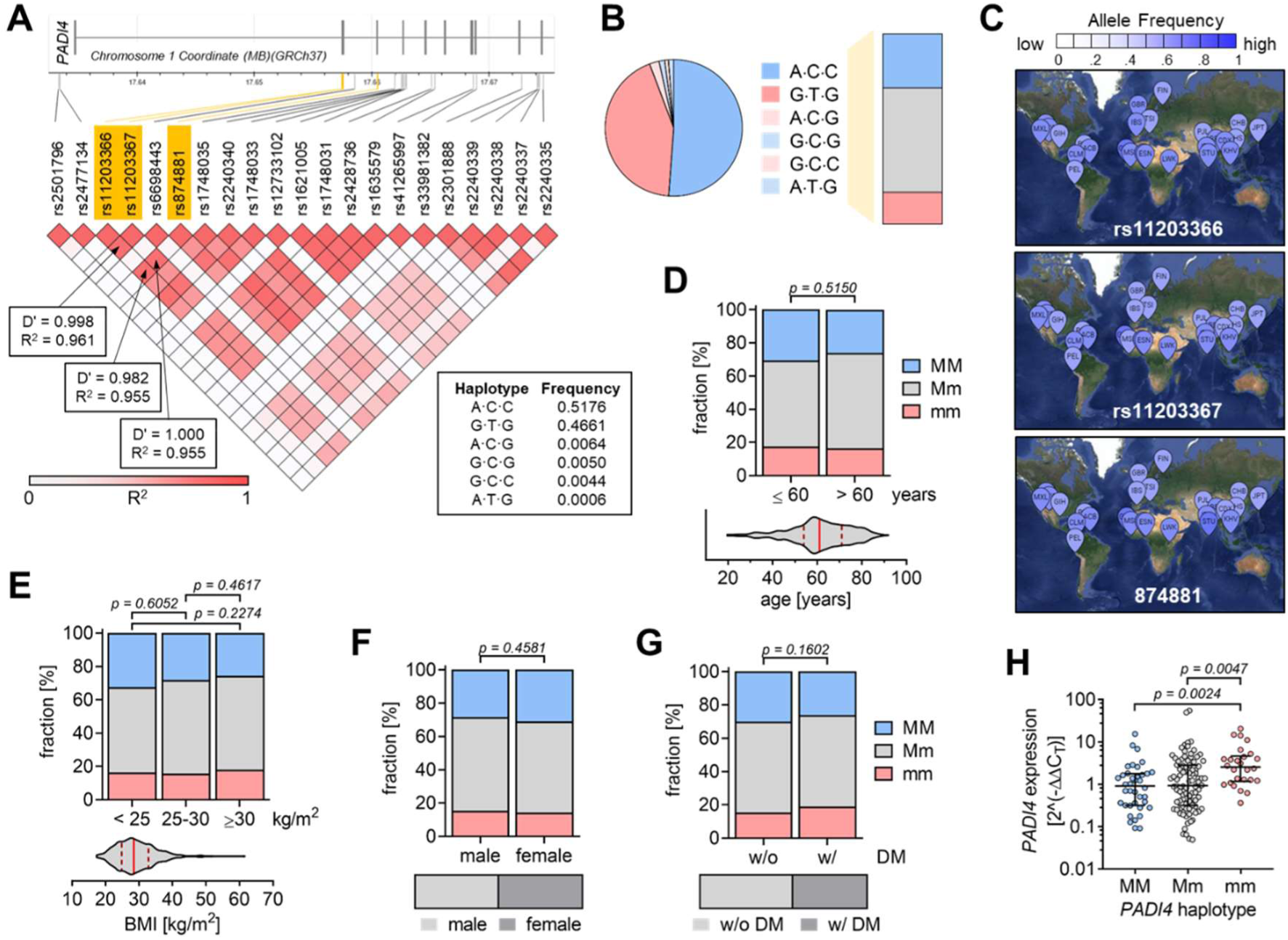
Distribution of the PADI4 haplotype within the study cohort. (A) SNPs within the coding region of the PADI4 gene were retrieved from the Ensemble Genome Browser (https://www.ensembl.org). SNPs with a minor allele frequency > 0·01 were fed into the LDmatrix Tool from the LDlink app (https://www.ldlink.nci.nih.gov). Shown are the obtained D’ (deviation of haplotype frequencies from expected values based on gene frequencies) and R^2^ (coefficient of determination). The three SNPs with the highest D’ and R^2^, located within the PADI4 exons, and coding for an amino acid shift were used in this study. The haplotype frequency of these three PADI4 SNPs (rs11203366, rs11203367, rs874881) was determined using the LDhap Tool from the LDlink program. (B) The corresponding PADI4 haplotype frequency of the study cohort was determined by ARMS-PCR. Major and minor haplotype (homozygous) are marked as MM and mm, respectively. Mm: heterozygous. (C) The worldwide allele frequency of the three PADI4 SNPs was determined using the LDpop Tool from the LDlink app. (D) Basal *PADI4* expression in circulating neutrophils (isolated prior to surgery) with different PADI4 haplotypes was determined by quantitative real-time PCR. Data are displayed as scatter plot and groups were compared by nonparametric Kruskal-Wallis test followed by Dunn’s multiple comparison test. Haplotype distribution was determined based on the patients’ (D) age, (E) body mass index (BMI), (F) sex, or (G) underlying disease (diabetes) and compared by chi-squared (χ^2^) test. (H) For all experiments a *p* < 0.05 was considered significant.

### PADI4 expression is increased in neutrophils with the PADI4 minor haplotype

A previous report stated that the PADI4 haplotype may affect the mRNA stability (*21*). Therefore, we tested if the basal neutrophilic PADI4 expression levels related to the PADI4 haplotype in our patients. Indeed, neutrophils from patients homozygous for the minor (mm) PADI4 haplotype showed significantly higher PADI4 mRNA levels (4.4 ± 5.5) than neutrophils from patients heterozygous (Mm: 2.0 ± 2.4) or homozygous for the major (MM: 1.1 ± 0.9) PADI4 haplotype (Figure 2H). Due to the high linkage disequilibrium of the three SNPs, similar was observed when investigating the effect of the individual SNPs on basal PADI4 expression levels (data not shown).

### Highest PADI4 expression in neutrophils from patients with DM developing complications

As increased neutrophilic PADI4 expression is supposed to affect wound healing (*16*), we related the PADI4 expression levels to the clinical outcome in our patients. PADI4 expression was overall higher in patients with DM when compared to patients without DM. While basal PADI4 expression levels did not significantly differ between patients without DM developing or not a complication, basal PADI4 expression was increased on average by 2.1-fold, 3.7-fold, and 3.1-fold, respectively, in patients with DM developing an infection (Figure 3A), delayed wound healing (DWH / Figure 3B), or delayed bone healing (DBH / Figure 3C). This also displayed in the corresponding receiver operating characteristic (ROC) curves, with areas under the curve (AUC) being significant for patients with DM (Infect: AUC = 0.6748, *p* = 0.0381 & DWH: AUC = 0.6949, *p* = 0.0216) but not for those without DM (Infect: AUC = 0.5209, *p* = 0.7162 & DWH: AUC = 0.5357, *p* = 0.5603) developing an infection or delayed wound healing (Figure 3D&E). For patients developing delayed bone healing a similar trend (diabetics: AUC = 0.6533, *p* = 0.1168 & controls: AUC = 0.5318, *p* = 0.6976) was observed (Figure 3F).

**Figure 3:**
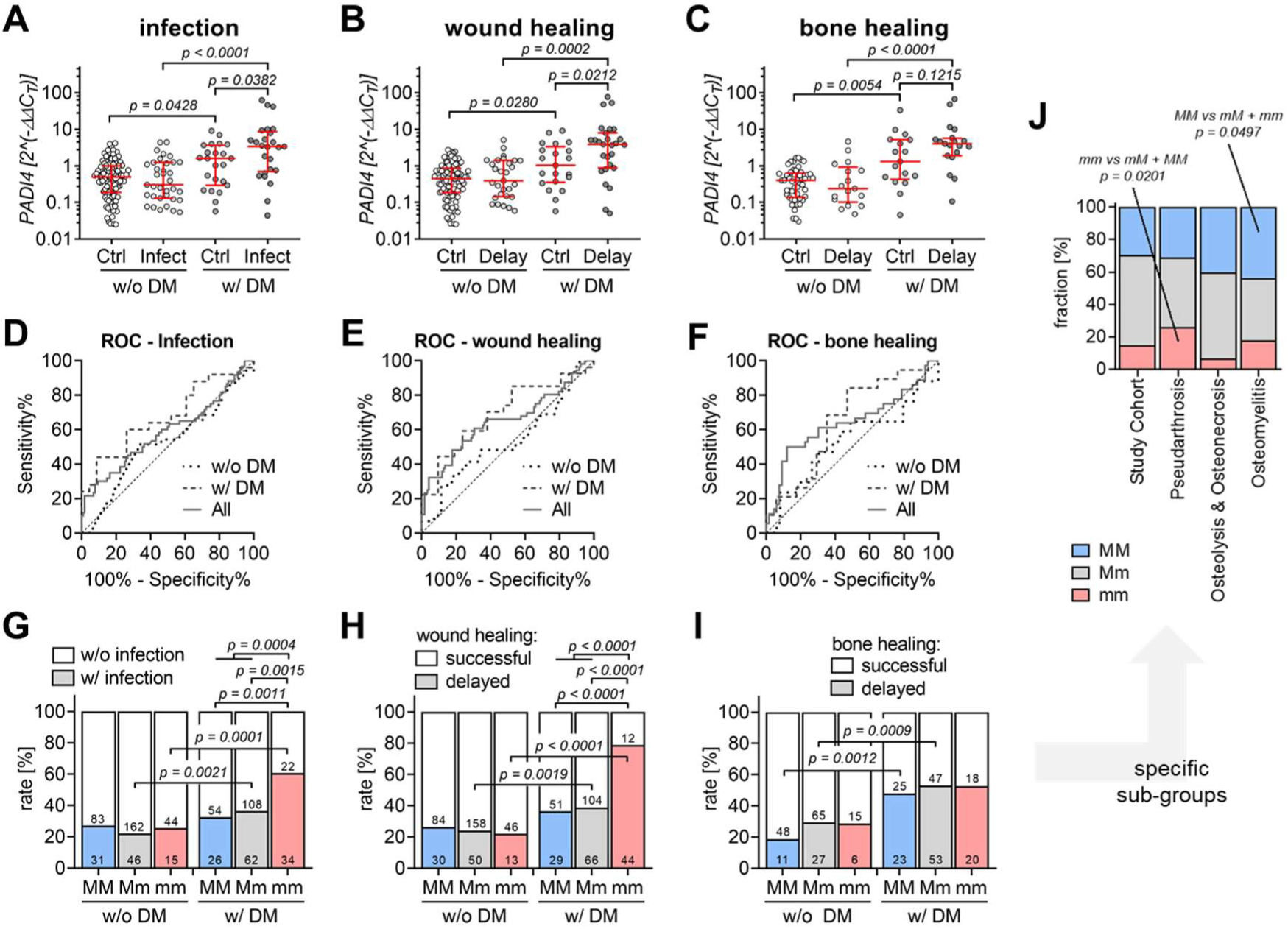
Association of the PADI4 haplotype with PADI4 expression and clinical outcome. (A) PADI4 expression in circulating neutrophils (isolated prior to surgery) was determined by qRT-PCR and related to the three PADI4 SNPs rs11203366, rs11203367, and rs874881, determined by ARMS-PCR. Neutrophilic PADI4 expression was then compared between patients with and without Diabetes mellitus (DM), that did or didn’t develop (B) infections, (C) delayed wound healing (DWH), and (D) delayed bone healing (DBH). Data are displayed in scatter plots and groups were compared by nonparametric Kruskal-Wallis test followed by Dunn’s multiple comparison test. The PADI4 expression levels were used to generate receiver operating characteristic (ROC) curve for the development of (E) infections, (F) delayed wound healing, and (G) delayed bone healing in control and diabetic patients. PADI4 haplotype distribution based on the patients’ underlying disease (controls vs. diabetics) and the clinical outcome, developing (H) infections, (I) delayed wound healing, and (J) delayed bone healing was compared by chi-squared (χ^2^) test. For all experiments a *p* < 0.05 was considered significant.

### Highest complication rates in patients with DM harboring the PADI4 minor haplotype

Based on the observed relation between neutrophilic PADI4 expression levels and both the PADI4 haplotype and the clinical outcome, the PADI4 haplotype was related to the clinical outcome of the patients. Overall, the herein analyzed complications were documented more often in diabetics than controls. In patients without DM, the rate of complications was comparable between patients harboring the major or minor PADI4 haplotype. However, among the patients with DM, those homozygous for the minor PADI4 haplotype developed significantly more infections (mm: 60.7%) than those heterozygous (Mm: 36.5%) or those homozygous for the major (MM: 32.5%) PADI4 haplotype (Figure 3G). The same holds for patients with DM and delayed wound healing – the frequency was highest in those homozygous for the minor PADI4 haplotype (mm: 78.6%), followed by those heterozygous (Mm: 38.8%) or homozygous for the major (MM: 36.3%) PADI4 haplotype (Figure 3H). For patients with DM and delayed bone healing no such trend was observed (Figure 3I). In order to investigate, if a specific pathology associated with delayed bone healing is related to the PADI4 haplotype, a subgroup analysis was performed that differentiated between specific bone-related complication, namely, pseudarthrosis (N = 54), osteomyelitis (N = 39), osteolysis and osteonecrosis (N = 30), independent of the DM-status. Interestingly, in patients with osteomyelitis, osteolysis and osteonecrosis there was an increase in the major PADI4 haplotype as compared to the entire study population. However, with infections present in pseudarthrosis and osteomyelitis, also an increase in minor PADI4 haplotype was observed (Figure 3J).

Single complication rates attributed to the PADI4 SNPs genotype or allele distribution in patients with and without DM are summarized in Table 1. In line with the PADI4 haplotype distribution the minor alleles of SNPs rs11203366 (A>G), rs11203367 (C>T), and rs874881 (C>G) strongly related to the development of infections and delayed wound healing in patients with DM.

**Table 1.** Genotype and allele distribution within the study.

|  |  | All (N w/o : N w/) |  |  | Ctrl (N w/o : N w/) |  |  | DM (N w/o : N w/) |  |  |
| --- | --- | --- | --- | --- | --- | --- | --- | --- | --- | --- |
|  |  | Infect | DWH | DBH | Infect | DWH | DBH | Infect | DWH | DBH |
| **rs11203366** |  |  |  |  |  |  |  |  |  |  |
| Genotypes | *p-value* | **0.0172** | **0.0005** | 0.0864 | 0.8894 | 0.7976 | 0.1919 | **0.0016** | **<0.0001** | 0.6415 |
|  | AA | 131 : 52 | 128 : 55 | 70 : 30 | 78 : 27 | 78 : 27 | 45 : 9 | 53 : 25 | 50 : 28 | 25 : 21 |
|  | AG | 277 : 114 | 270 : 121 | 115 : 84 | 168 : 51 | 165 : 54 | 68 : 29 | 109 : 63 | 105 : 67 | 47 : 55 |
|  | GG | 65 : 48 | 57 : 56 | 33 : 26 | 43 : 14 | 45 : 12 | 15 : 6 | 22 : 34 | 12 : 44 | 18 : 20 |
| Alleles | *p-value* | **0.0370** | **0.0047** | 0.0635 | 0.8099 | 0.5767 | 0.1706 | **0.0033** | **<0.0001** | 0.5158 |
|  | A | 539 : 218 | 526 : 231 | 255 : 144 | 324 : 105 | 321 : 108 | 158 : 47 | 215 : 113 | 205 : 123 | 97 : 97 |
|  | G | 407 : 210 | 384 : 233 | 181 : 136 | 254 : 79 | 255 : 78 | 98 : 41 | 153 : 131 | 129 : 155 | 83 : 95 |
| **rs11203367** |  |  |  |  |  |  |  |  |  |  |
| Genotypes | *p-value* | **0.0145** | **0.0004** | 0.1119 | 0.6937 | 0.6574 | 0.2789 | **0.0018** | **<0.0001** | 0.8392 |
|  | CC | 139 : 57 | 135 : 61 | 72 : 32 | 86 : 31 | 85 : 32 | 49 : 11 | 53 : 26 | 50 : 29 | 23 : 21 |
|  | CT | 268 : 108 | 262 : 114 | 112 : 81 | 159 : 46 | 157 : 48 | 64 : 27 | 109 : 62 | 105 : 66 | 48 : 54 |
|  | TT | 66 : 49 | 58 : 57 | 34 : 27 | 44 : 15 | 46 : 13 | 15 : 6 | 22 : 34 | 12 : 44 | 19 : 21 |
| Alleles | *p-value* | **0.0432** | **0.0073** | 0.0683 | 0.7326 | 0.4086 | 0.2066 | **0.0045** | **<0.0001** | 0.6683 |
|  | C | 546 : 222 | 532 : 236 | 256 : 145 | 331 : 108 | 327 : 112 | 162 : 49 | 215 : 114 | 205 : 124 | 94 : 96 |
|  | T | 400 : 206 | 378 : 228 | 180 : 135 | 247 : 76 | 249 : 74 | 94 : 39 | 153 : 130 | 129 : 154 | 86 : 96 |
| **rs874881** |  |  |  |  |  |  |  |  |  |  |
| Genotypes | *p-value* | **0.0468** | **0.0009** | 0.2292 | 0.5401 | 0.5879 | 0.3927 | **0.0018** | **<0.0001** | 0.8392 |
|  | CC | 125 : 55 | 123 : 57 | 68 : 32 | 77 : 30 | 77 : 30 | 45 : 11 | 48 : 25 | 46 : 27 | 23 : 21 |
|  | CG | 270 : 107 | 264 : 113 | 112 : 81 | 158 : 46 | 157 : 47 | 64 : 27 | 112 : 61 | 107 : 66 | 48 : 54 |
|  | GG | 78 : 52 | 68 : 62 | 38 : 27 | 54 : 16 | 54 : 16 | 19 : 6 | 24 : 36 | 14 : 46 | 19 : 21 |
| Alleles | *p-value* | 0.1418 | **0.0123** | 0.1812 | 0.3889 | 0.3998 | 0.4616 | **0.0075** | **<0.0001** | 0.6683 |
|  | C | 520 : 217 | 510 : 227 | 248 : 145 | 312 : 106 | 311 : 107 | 154 : 49 | 208 : 111 | 199 : 120 | 94 : 96 |
|  | G | 426 : 211 | 400 : 237 | 188 : 135 | 266 : 78 | 265 : 79 | 102 : 39 | 160 : 133 | 135 : 158 | 86 : 96 |

*P-values* are given for the comparison of each MM vs. mm (genotypes) and M vs. m (alleles). Groups were compared by chi-squared ( $\chi^2$ ) test. DWH = delayed wound healing; DBH = delayed bone healing

### PADI4 haplotype was associated with PADI4 protein levels and PADI4 activity

As the PADI4 haplotype was related to the PADI4 gene expression levels, we tested their effect on PADI4 protein levels in neutrophils isolated from healthy donors homozygous for the PADI4 major or minor haplotype (Figure 4A). Neutrophils with the PADI4 minor haplotype had 2.0-fold higher basal PADI4 levels than neutrophils with the PADI4 major haplotype. As a marker for PADI4 activity the levels of citrullinated histone H3 (cit-H3) were quantified in these cells. Although, basal cit-H3 levels were very low, their levels were 3.5-fold higher in neutrophils with the PADI4 minor haplotype when compared to neutrophils with the PADI4 major haplotype (Figure 4B&C). As additional neutrophil markers protein levels of neutrophil elastase (ELANE) and myeloperoxidase (MPO) were quantified. While ELANE levels were not significantly affected by the PADI4 haplotype, basal MPO levels were 1.7-fold higher in neutrophils with the PADI4 major haplotype when compared to neutrophils with the PADI4 minor haplotype (Figure 4D&E).

**Figure 4:**
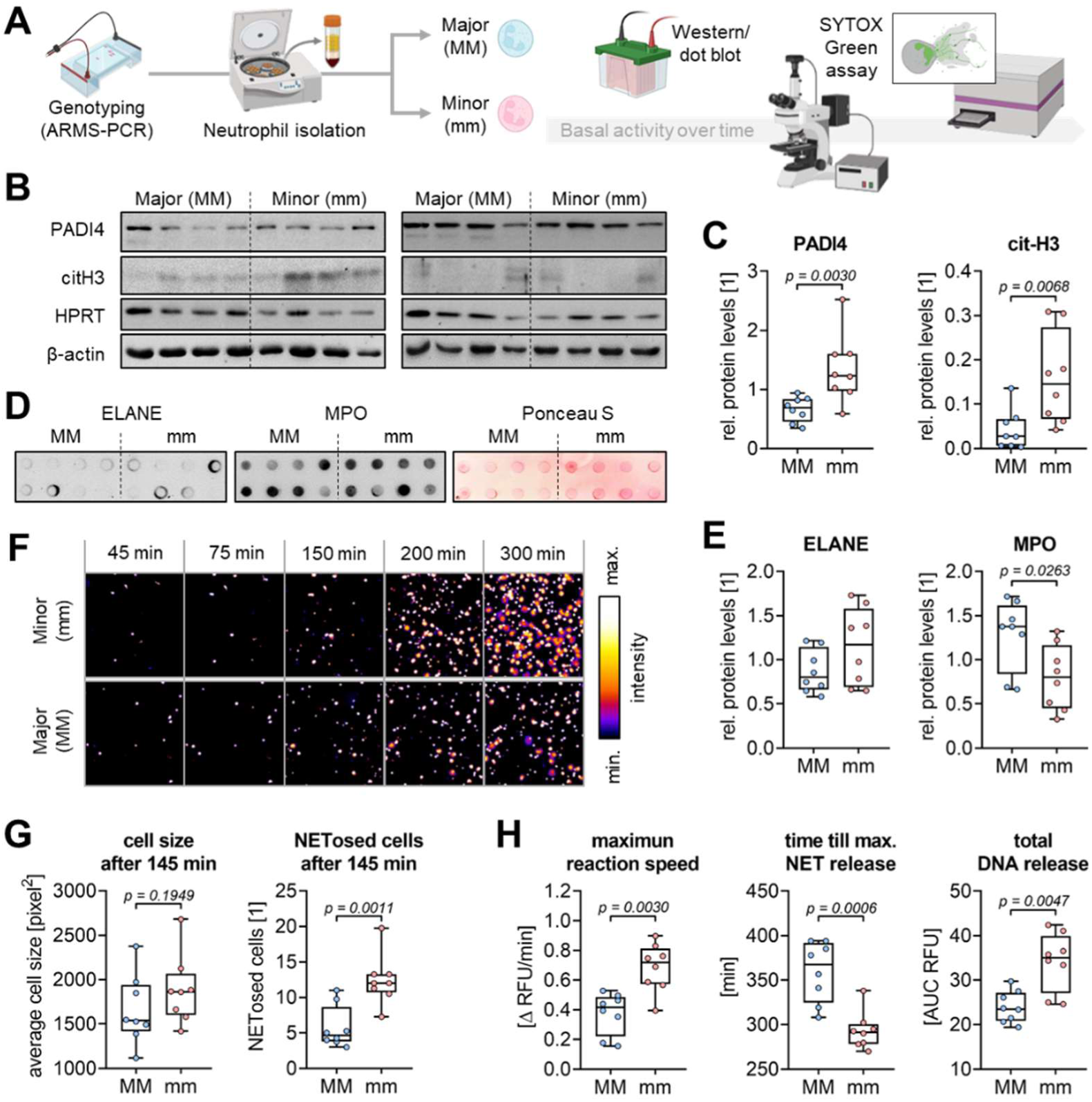
Basal NET formation in neutrophils with different PADI4 haplotype. (A) Schematic overview of the experimental setup – created with icons from BioRender https://www.biorender.com/. Freshly isolated neutrophils from 16 healthy donors homozygous for the major (MM: N = 8) or minor (mm: N = 8) PADI4 haplotype were examined for basal neutrophil activation. (B) Western blot images for PADI4 and citrullinated Histone H3 (citH3). (C) Western blot signals for PADI4 and citH3 were quantified by ImageJ software (each membrane with n = 3 exposure times). HPRT and β-actin were used as loading controls. (D) Dot blot images for neutrophil elastase (ELANE) and myeloperoxidase (MPO). (E) Dot blot signals for ELANE and MPO were quantified by ImageJ software (each membrane with n = 3 exposure times). Ponceau S staining was used as loading control. (F) Representative time-laps images of the performed SYTOX^TM^ Green Assay. SYTOX^TM^ Green signal intensities were visualized in ImageJ using “fire” pseudocolor. (G) Morphologic analyses (*22*) were performed to detect the average cell size and the number of NETosed cells after 145 min. (H) Curve fitting was done to determine the maximum NET release rate, represented by the strongest increase (slope) in fluorescence, and the time point of maximum NET release, represented by the inflection point of the slope. The total DNA released was determined by the area under the curve (AUC). Data are displayed in box plots with individual data points, median, and interquartile range. Data between two groups were compared by Mann-Whitney U-tests. A *p* < 0.05 was considered significant and is shown in the graphs.

### Increased basal NET formation in neutrophils homozygous for the PADI4 minor haplotype

The same neutrophils were analyzed for their potential to form NETs by SYTOX^TM^ Green Assay. Time-laps fluorescent images showed more basal NET formation in neutrophils with the PADI4 minor haplotype when compared to neutrophils with the PADI4 major haplotype (Figure 4F). Image analysis confirmed an increased cell size and higher amount of NETosed cells (Figure 4G) 145 min after isolation. Fluorescent signals quantified with an Omega plate reader were used for curve fitting. Basal NET formation occurred more rapidly (1.7-fold with Δ 76 min) in neutrophils homozygous for the PADI4 minor haplotype when compared to neutrophils homozygous for the PADI4 major haplotype. Further, neutrophils with the PADI4 minor haplotype released 1.5-fold more DNA than neutrophils with the PADI4 major haplotype (Figure 4H).

### PADI4 minor haplotype sensitized neutrophils towards NETosis stimuli

The freshly isolated neutrophils from healthy volunteers homozygous for the PADI4 major or minor haplotype were then stimulated with 100 nM PMA or 4 µM CI, in order to induce NETosis. Within 5 h following stimulation different aspects of NET formation were documented (Figure 5A). Cell attachment and activation were detected by bioimpedance measurement (Figure 5B). Directly after stimulation, the group with the PADI4 minor haplotype showed a more rapid increase in cell index (cell attachment: PMA: 2.0-fold and Δ 17 min; CI: 1.9-fold) than the group with the PADI4 major haplotype. The reaction speed following CI stimulation was so quick that no difference between the two groups could be detected. Later, when NET release predominated, the cell index decreased more rapidly (PMA: 2.8-fold; CI: 1.7-fold) in the group with the PADI4 minor haplotype when compared to the group with the PADI4 major haplotype (Figure 5C). As another aspect of NETosis, the DNA release was quantified by SYTOX^TM^ Green Assay (Figure 5D). Neutrophils homozygous for the PADI4 minor haplotype more rapidly released (PMA: 1.3-fold with Δ 18 min; CI: 1.7-fold) a larger amount of DNA than neutrophils homozygous for the PADI4 major haplotype (Figure 5E). To verify that necrotic cells did not interfere with the SYTOX^TM^ Green Assay, time-resolved fluorescent images were analyzed. The representative compilation shows, that following stimulation neutrophils with the PADI4 minor haplotype showed more NETosed cells (diffuse fluorescence) than neutrophils with the PADI4 major haplotype. Noteworthy, stimulation with CI induced not only NETosis but also necrosis (condensed fluorescence) in the neutrophils (Figure 5F). Automated image analyses confirmed this observation – in PMA stimulated neutrophils, the average cells size and number of NETosed cells were significantly higher in the group with the PADI4 minor haplotype (1.3-fold and 1.5-fold, respectively) when compared to the group with the PADI4 major haplotype (Figure 5G).

**Figure 5:**
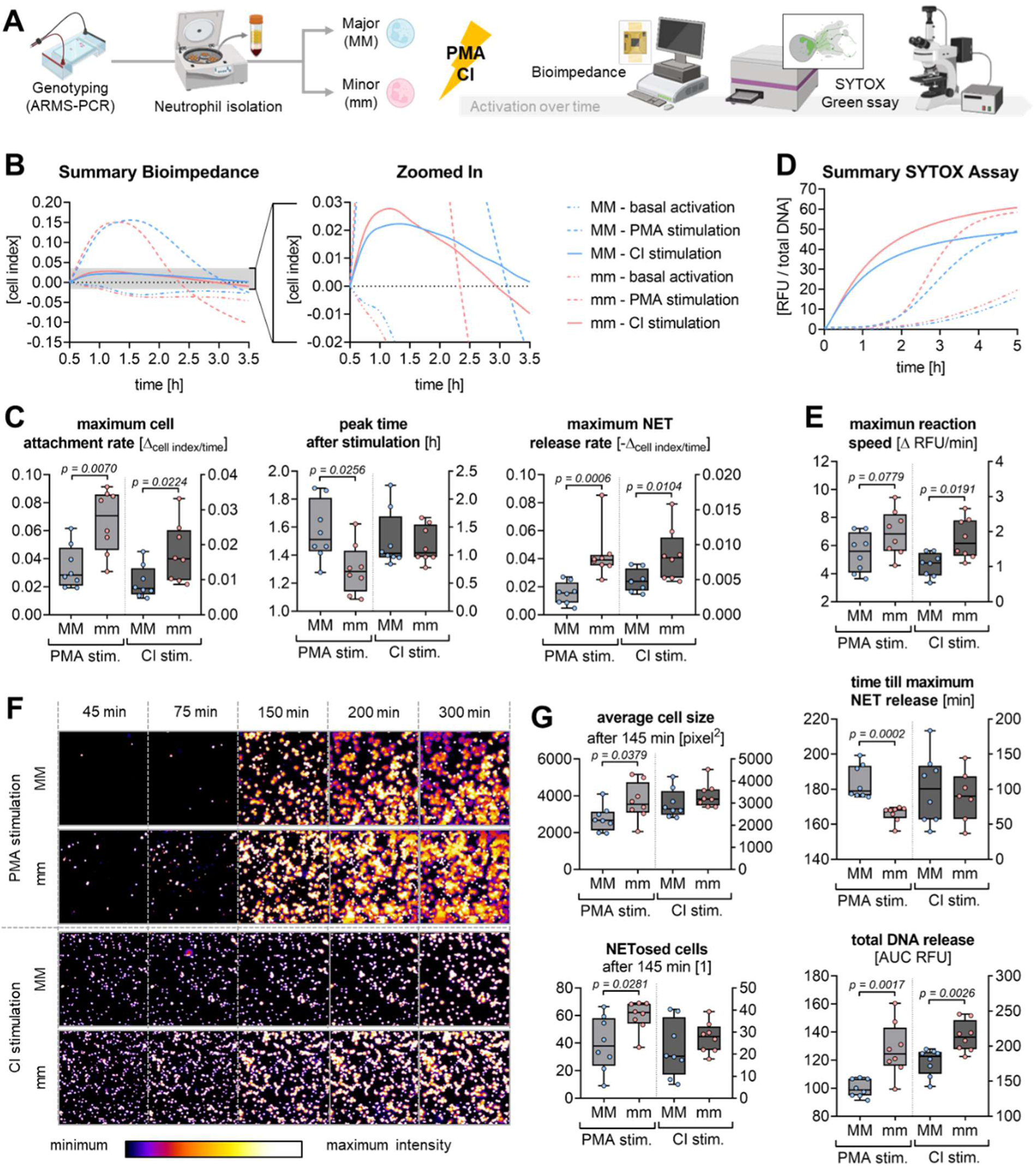
PMA- and CI-induced NET formation in neutrophils with different PADI4 haplotype. (A) Schematic overview of the experimental setup – created with icons from BioRender https://www.biorender.com/. Neutrophils from 16 healthy donors homozygous for the major (MM) or minor (mm) PADI4 haplotype were examined for neutrophil activation upon stimulation with 100 nM Phorbol 12-myristate 13-acetate (PMA) or 4 µM calcium ionophore A23187 (CI), respectively. Bioimpedance measurement [cell index], using the xCelligence RTCA eSight device, was used to identify early events in neutrophil activation. (A) Curve fitting for the average cell index measured (N = 8, N = 4 per group). (B) The maximum cell attachment rate was determined by the strongest increase (slope) in cell index. (C) The peak cell index represents the time point when NET release starts to superpose neutrophil attachment. (D) The maximum NET release rate is presented by the strongest decrease (slope) in cell index. In the same cells NET formation / DNA release was quantified by SYTOX^TM^ Green Assay. (E) Curve fitting for the measured fluorescent intensities (N = 8, N = 3 per group). (F) The maximum NET release rate was determined by the strongest increase (slope) in fluorescence. (G) The time point of maximum NET release is represented by the inflection point of the slope. (H) The total DNA released was determined by the area under the curve (AUC). Using the xCelligence RTCA eSight device allowed online detection of fluorescent images from the SYTOX^TM^ Green Assay. (I) Representative time-laps images of the performed SYTOX^TM^ Green Assay with fluorescent intensities visualized in ImageJ using “fire” pseudocolor. Morphologic analyses (*22*) were performed to detect (J) the average cell size and (K) the number of NETosed cells after 145 min. Data are displayed in box plots with individual data points, median, and interquartile range. Data between two groups were compared by Mann-Whitney U-tests. A p < 0.05 was considered significant and is shown in the graphs.

## Discussion

This study is the first to establish a link between the human PADI4 haplotype (SNPs: rs11203366, rs11203367, rs874881) and wound healing complications in patients with DM undergoing orthopedic and trauma surgery. Furthermore, this functional PADI4 haplotype was linked to neutrophilic PADI4 expression and influenced the neutrophils’ ability to form NETs.

Tissue trauma, such as a wound or fracture, leads to release of mitochondrial DNA, which then stimulates neutrophils to form NETs at the site of injury (*23*). Under normal conditions, this is thought to prevent pathogens from colonizing the wound or entering the bloodstream to develop a sepsis. Under diabetic conditions, however, NET accumulation was observed in the wounds, which impaired the healing process (*7*). The resulting increase in circulating NET markers remains even after chronic DFU have developed, and may predict the healing outcome (*12, 15*). In our patients, however, the development of chronic wounds should be prevented. Considering that the accumulated NETs cause damage to the tissue, measuring the amount of NETs formed may not be a suitable approach as it leaves little window of opportunity to adjust treatment. In the diabetic mouse model neutrophilic overexpression of PADI4 was identified as the key inducer for the excessive NET formation (*7*). In line with this, pre-surgical neutrophilic PADI4 levels were significantly increased in our trauma patients with DM, especially in those who later experienced delayed wound healing and/or developed a wound infection. *In vitro*, forced overexpression of PADI4 was reported to induce histone hypercitrullination leading to heterochromatin decondensation and chromatin unfolding (*24*), as well as oxidative burst (*25*), which in case of neutrophils sensitizes the cells for NET stimuli and may even induce spontaneous NET formation. This may explain, why in our experiments both basal and PMA-induced NET formation was increased in neutrophils with increased PADI4 expression.

In our patients, pre-surgical PADI4 expression associated not only with the clinical outcome, but also with the investigated PADI4 haplotype – with highest PADI4 expression observed in neutrophils coding for the G T G alleles (minor variant). This is partly in contrast to patients with septic shock, in whom increased PADI4 levels were positively associated to mortality but not with PADI4 SNP rs11203366 (*26*), however, this might be due to the small sample size in the mortality group (N = 41). A study characterizing the PADI4 protein structure proposed, that the base changes in our investigated PADI4 haplotype may alter the PADI4 mRNA stability with prolonged mRNA stability for the minor variant (*21*). This is supporting our findings, that neutrophils with the PADI4 minor haplotype had higher PADI4 mRNA and protein levels. This in turn related with a basal increase in citH3, suggesting that the PADI4 minor haplotype sensitizes neutrophils to release NETs. Indeed, PMA- or CI-induced NET formation was faster and stronger in neutrophils with the PADI4 minor haplotype than in neutrophils with the PADI4 major haplotype.

In line with the premise that excessive NET formation in wound tissue is associated with disturbed wound healing in patients with DM, highest PADI4 expression and higher frequency of PADI4 minor haplotype was found in neutrophils of patients who later developed a wound healing disorder. Interestingly, the increased complication rate associated with the PADI4 minor haplotype could only be shown in patients with DM, suggesting that DM provides additional NET stimuli. It was proposed that increased glucose levels sensitize neutrophils to form NETs (*27*), more specifically, that exogenous glucose and glycolysis promotes the actual NET release but not the initial chromatin decondensation (*28*). Another study proposed that hyperglycemia supports just the latter by nuclear accumulation of acetyl-CoA, which acts as substrate for histone acetyltransferases (*29*). Yet another study suggested that not the glucose itself, but increased homocysteine levels in DM are the actual trigger for NET formation (*30*). It is questionable if both factors can be discussed independently, as homocysteine levels are positively associated with insulin resistance, dyslipidemia, and poor glucose control (*31*). Both have in common that they induce cellular stress, which increases intracellular calcium levels, a process involving gasdermine E (*32, 33*). The increase in intracellular calcium may than act as a co-factor for the PADI4 enzyme (*34*), but also independently stimulate NET formation as observed for stimulation with calcium-ionophore (*35*).

Hyperglycemia is well recognized as a normal metabolic stress response. Therefore, it should be considered that our study cohort consists of trauma patients. In such patients’ blood glucose control is aggravated by the stress induced by a trauma and following surgery, resulting in frequent hyperglycemic episodes (*36*). The resulting elevation in basal PADI4 expression might explain, why NET markers are stronger increased in patients with DM following trauma, but also offers the opportunity for an intensified blood glucose control upon hospital admission. First hints are provided from studies with critically-ill patients (ΣN > 7,000) in the intensive care unit (ICU) (*37, 38*). In these patients, strict blood glucose control by intensified insulin treatment (IIT) reduced morbidity and mortality, with the consequence of faster weaning of the mechanical ventilation, discharge from the ICU and hospital. However, a subgroup analysis showed that only trauma patients and patients receiving corticosteroid therapy may have an advantage in survival upon IIT (*40*). Furthermore, strict glycemic control was shown to significantly reduce (50-60%) the incidence of nosocomial infections in surgical ICU patients (*39*).

Our data clearly link the PADI4 haplotype with the neutrophilic PADI4 expression in humans, which in turn sensitizes the cells to form NETs. Therefore, genotyping for the PADI4 haplotype might become a relevant screening tool in the future, not only to identify patients with DM at increased risk of developing wound healing disorders but also for other pathologies, *e.g.*, RA, sepsis, deep vein thrombosis, multiple sclerosis, *etc.*, where excessive NET formation and more precisely dysregulated PADI4 activity has been shown to aggravate disease progression (*16*). This is of special importance as such a haplotype screening can be done at the initial diagnosis of the disease, which allows for early intervention. Considering the pathologic role of excessive NET formation could identify novel therapeutic options for such diseases, *e.g.*, enzymatic digestion of he formed NETs (*7*) or ir-/reversible PADI4 inhibitors, *e.g.*, GSK199, GSK484, TDCA, TDFA, JBI-589, F- or Cl-amidine (*34, 41*). However, treatment strategies should involve suppression of excessive NET formation not total depletion of NETs, as this might increase the risk for infections and sepsis (*42, 43*).

## Conclusion

In summary, we here provide first evidence that the PADI4 genetic variation influences neutrophil behavior and clinical outcomes. The PADI4 minor haplotype sensitized neutrophils toward NET formation and was associated with an increased complication rate in patients with DM following orthopedic or trauma surgery. PADI4 haplotyping may provide a clinically actionable biomarker to identify patients with DM at high risk for wound healing and infectious complications early upon hospital admission, enabling closer monitoring and/or personalized treatment to prevent complications.

## Methods

### Study cohort and sampling

Between September 2018 and February 2022, 687 patients were included in the study by giving their written informed consent (666/2018BO2). During recruitment there was no discrimination by sex, resulting in a sex distribution of 57.1% males (N = 392) and 42.9% females (N = 295). From each individual, swabs of the mucous membrane or EDTA-venous blood were collected. Information about the clinical outcome was independently collected by two physicians in a blinded manner. Disturbed wound healing, delayed bone healing, and post-surgical infections were rated as complications, both as result of a surgery or as cause for a revision surgery. Noteworthy, bone healing was not considered in patients receiving a total joint arthroplasty. Clinical data were merged with the laboratory data only when data acquisition was finished and verified by a second investigator.

### Isolation of neutrophils

Neutrophils were isolated from EDTA-venous blood by density gradient centrifugation, as described in (*44*). Isolated neutrophils were quantified by Trypan Blue exclusion method, then diluted to obtain a density of 1 Mio neutrophils/mL (concentrated cell suspension).

### qRT-PCR

Total mRNA from freshly isolated neutrophils was isolated by phenol-chloroform extraction and photometrically quantified. Total mRNA was converted into cDNA using the first-strand cDNA synthesis kit (ThermoFisher Scientific, Sindelfingen, GER). Quantitative RT-PCRs were performed using the Green Master Mix (2X) High Rox (Biozym, Hessisch Oldendorf, GER) and the StepOnePlus^TM^ qPCR cycler. Specificity of the qPCR reactions was checked by melting curve analyses. Relative PADI4 expression levels were calculated using the ΔΔC_T_ method, normalized to *EF1α* and *RPL13a*, which proved to be most stable (GeNorm, NormFinder, Best Keeper, comparative ΔCt method) between individuals (*45*). PCR conditions are summarized in Table 2.

**Table 2.**
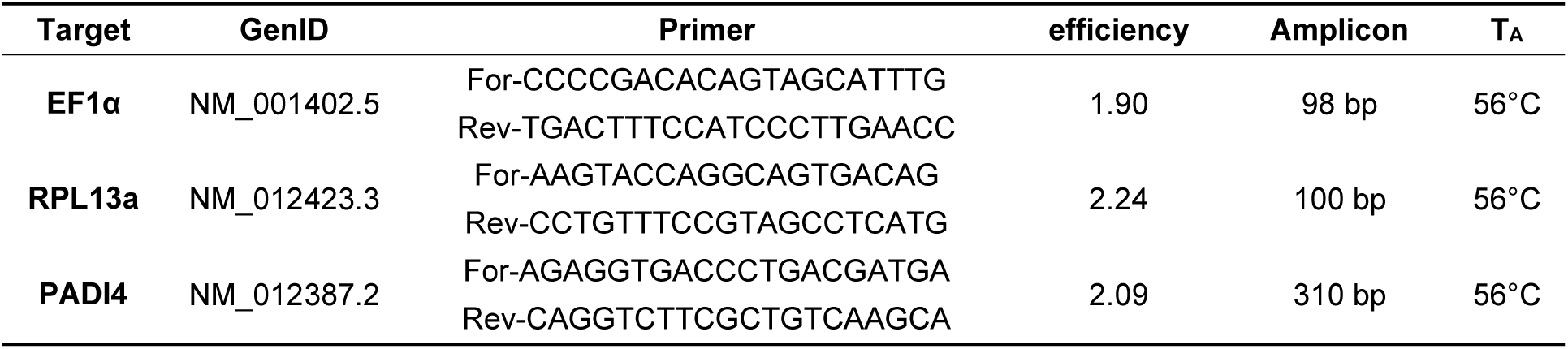
Detailed information on primers (designed with primer blast) and the corresponding optimized qPCR conditions.

### Amplification Refractory Mutation System (ARMS) PCR

DNA was isolated from residual leukocytes obtained from neutrophil isolation or swabs of the mucous membrane, as described in (*46*). SNPs were characterized by ARMS-PCR (*46*), using 3 µL of DNA (50-100 ng/µL), a mix of 4 primers (Table 3), and the Red HS Taq (2X) Master Mix (Biozym). The amplified DNA was separated by electrophoresis using a 2% agarose gel and visualized with ethidium bromide.

**Table 3.**
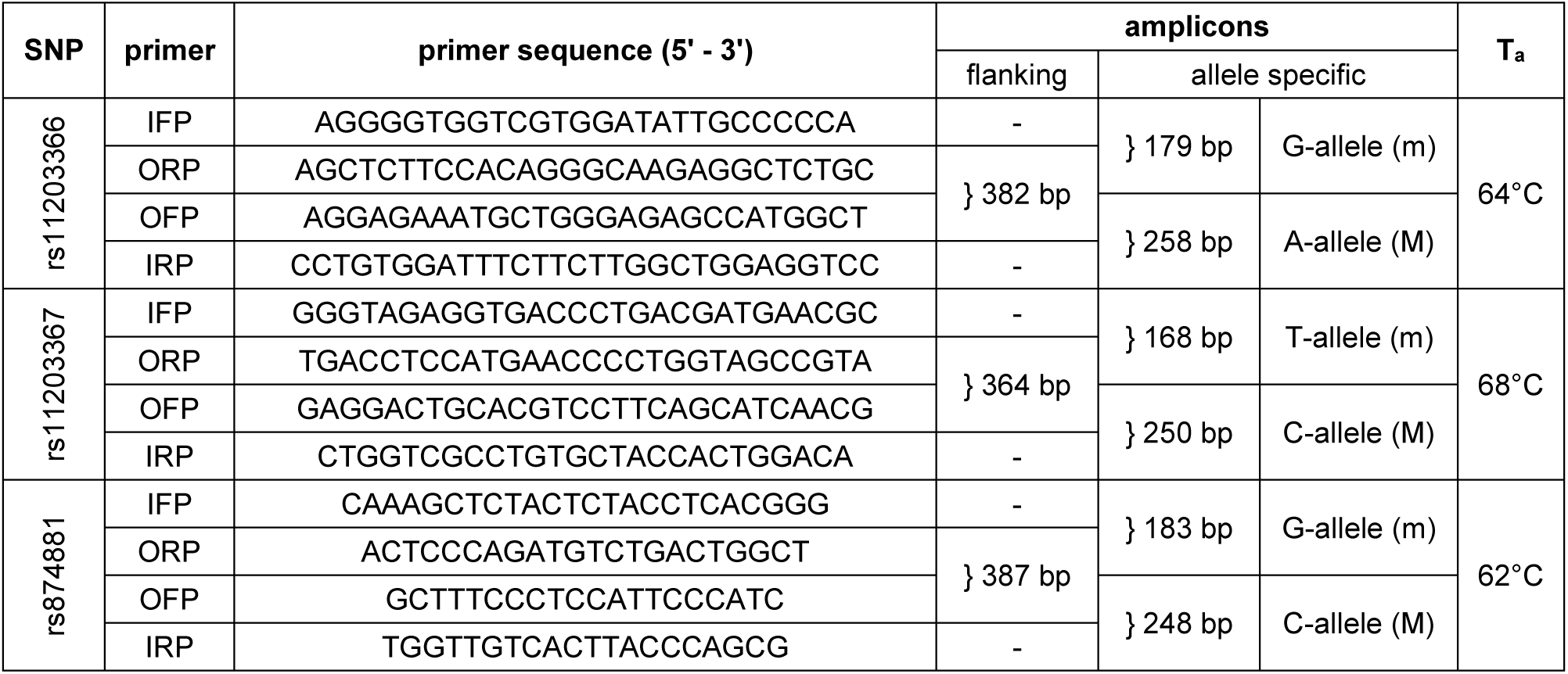
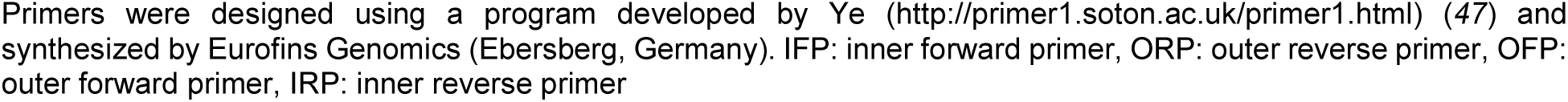
Primer sequences and ARMS-PCR conditions.

### Western blot and dot blot

Neutrophils were lyzed with ice-cold RIPA buffer with protease- and phosphatase inhibitors. For Western blot 35 µg total protein was separated by SDS-PAGE and transferred to nitrocellulose membranes. For dot blot 35 µg total protein was directly applied to nitrocellulose membranes under vacuum using a dot blotter (Carl Roth, Karlsruhe, GER). Protein transfer to membraned was confirmed by Ponceau S staining. Unspecific binding sites were blocked with 5% BSA for 1 h. After overnight incubation with primary antibodies at +4°C (PADI4: sc-365369, HPRT: sc-376938, ELANE: sc-55548, MPO: sc-52707 - SantaCruz Biotechnology, Heidelberg, GER); Cit-H3: ab5103 (abcam, Cambridge, GBR); β-Actin: #4970 (Cell Signaling Technology, Danver, MA, USA)), membranes were incubated with the corresponding peroxidase-labeled secondary antibodies for 2 h. Enhanced chemiluminescence solution was added and the resulting signals were detected with a CCD camera and quantified with ImageJ.

### Stimulation of neutrophils

For all functional assays, neutrophils were stimulated with PMA (Phorbol 12-myristate 13-acetate) or calcium ionophore A23187 (CI). Solutions were prepared as 1.25-fold of the final concentration to allow measurement of blank (80 µL) and thereafter addition of concentrated cell suspension (20 µL) to receive a final concentration of 2*10^5^ neutrophils/mL (*44*).

### SYTOX^TM^ Green Assay

1.25 µM SYTOX^TM^ Green was added to the stimulation solutions before the addition of neutrophils. Cells were lysed with 1% Triton-X-100 (Carl Roth, Karlsruhe, Germany) for normalization and better comparability of donors. Fluorescence (λ_ex_=485 nm & λ_em_=520 nm) was measured every 30 min with the Omega Plate Reader (BMG Lab-tech) at 37°C and 5% CO_2_. Curve fitting of the resulting XY diagrams with GraphPad Prism included; (1) the maximum slope, which equals the maximum NET release rate; (2) the inflection point of the slope, which defines the time till maximum NET release; and (3) the area under the curve (AUC), which represents the total DNA released (*44*).

### Bioimpedance measurements and real-time fluorescent imaging

The xCelligence RTCA eSight device (Omni Life Sciences), placed in an incubator at 37°C with 5% CO_2_ and humidified atmosphere, was used to measure bioimpedance [cell index] (*44*). Neutrophils were stimulated in xCelligence measurement plates containing SYTOX^TM^ Green for real-time fluorescent imaging. After blank measurements, neutrophils were added and cell index was measured at least every 15 min for a period of 6 h. Fluorescence pictures were captured at least every 30 min. Curve fitting of the exported normalized data with GraphPad Prism included; (1) the maximum positive slope, which equals the maximum cell attachment rate; (2) the peak time after stimulation, which represents the time point when cell attachment and NET release are balanced; and (3) the minimum negative slope, which represents the maximum NET release rate. Real-time fluorescent images were exported as single tiff images and compiled in ImageJ: SYTOX^TM^ Green signal intensities were visualized in “fire” pseudocolor. Images of the 145 min time point (one image per well of the GFP channel) were subjected to automatic counting (particles >1,000 pixel^2^), using an auto local threshold (Bernsen, radius 30) and watershed separation on the respective binary images.

### Statistics

Patient data are displayed in violin plots, visually confirming a Gaussian distribution (skewness and kurtosis between −1 and 1). Data comparison was done by ordinary one-way ANOVA followed by Tukey’s multiple comparison test. Complication rates in the different groups were compared by chi-squared (χ^2^) test. *In vitro* experiments were performed with neutrophils from 8 donors per group (N = 8) with at least three technical replicates (n ≥ 3). Data are represented as box plots with individual measurement points, median, and interquartile range. Given the smaller sample size, Gaussian distribution cannot be assumed for the *in vitro* experiments, thus, data were compared by Mann-Whitney U-tests. For all experiments a *p* < 0.05 was considered significant. GraphPad Prism 8.0.1 was used to perform statistical analysis and generate graphs.

### Study approval

The study was approved by the ethics committee of the University Hospital Tübingen (ethical vote 666/2018BO2, approved 20.09.2018).

### Author contributions

S.E. – conceptualization of the study; funding acquisition; methodology and investigations; formal analysis and validation of the laboratory data; data visualization; writing of the manuscript draft. P.H. & C.I. – patient recruitment and retrieval of patient data; validation of the clinical (trauma) data. C.L. & J.M. – methodology and investigations. P.-G.A. & R.L. – resources and patient data; validation of the clinical (diabetes) data. G.B. – statistic counseling. S.P. & A.F. – recourses, validation of the clinical (diabetes) data. H.B., M.F.R. & T.H. – resources and patient data. A.K.N. – resources; supervision; validation of the laboratory data. All authors have reviewed and edited the manuscript and agreed to its published version.

## Data Availability

All data produced in the present work are contained in the manuscript.

